# *STK11* and *KEAP1* Mutations as Prognostic Biomarkers in an Observational Real-World Lung Adenocarcinoma Cohort

**DOI:** 10.1101/2020.01.23.20017566

**Authors:** Simon Papillon-Cavanagh, Parul Doshi, Radu Dobrin, Joseph Szustakowski, Alice M. Walsh

## Abstract

**Importance:** Understanding the mechanisms of primary resistance to immune checkpoint blockade therapy is of paramount importance for treatment selection. Somatic mutations in *STK11* and *KEAP1*, frequently co-mutated in nonsquamous non–small cell lung cancer, have been associated with poor response to immune checkpoint blockade. However, previous reports lack non–immune checkpoint blockade controls needed to properly ascertain the predictive nature of those biomarkers.

**Objective:** To evaluate the predictive vs prognostic effect of *STK11* or *KEAP1* mutations across different treatment classes in nonsquamous non–small cell lung cancer.

**Design:** A retrospective, real-world data cohort from the Flatiron Health network linked with genetic testing from Foundation Medicine, from January 1, 2011, through December 31, 2018.

**Setting:** Multicenter, including academic and community practices.

**Participants:** Patients diagnosed with stage IIIB, IIIC, IVA, or IVB nonsquamous non–small cell lung cancer who initiated first-line treatment within 90 days after diagnosis.

**Main Outcomes and Measures:** Real-world, progression-free survival and overall survival calculated from time of initiation of first-line treatment.

**Results:** We analyzed clinical and mutational data for 2276 patients with advanced, nonsquamous non–small cell lung cancer (mean age at advanced diagnosis, 66.3 years [SD 10.3], 54.4% female, 80.1% with a history of smoking), including patients treated with anti–programmed death-1/anti–programmed death ligand 1 inhibitors at first line (n = 574). Mutations in *STK11* or *KEAP1* were associated with poor outcomes across multiple therapeutic classes and were not specifically associated with poor outcomes in immune checkpoint blockade cohorts. There was no observable interaction between *STK11* mutations and anti–programmed death-1/anti–programmed death ligand 1 treatment on real-world, progression-free survival (HR, 1.05; 95% CI, 0.76-1.44; *P* = .785) or overall survival (HR, 1.13; 95% CI, 0.76-1.67; *P* = .540). Similarly, there was no observable interaction between *KEAP1* on real-world, progression-free survival (HR, 0.93; 95% CI, 0.67-1.28; *P* = .653) or overall survival (HR, 0.98; 95% CI, 0.66-1.45; *P* = .913). Results were consistent in *KRAS*-mutated patients.

**Conclusion and Relevance:** Our results show that *STK11*-*KEAP1* mutations are prognostic, not predictive, biomarkers for anti–programmed death-1/anti–programmed death ligand 1 therapy.

**Key Points:** *Question:* Are loss-of-function somatic mutations in *STK11* and *KEAP1* predictive of response to immune checkpoint blockade or simply prognostic?

*Findings:* In this observational real-world cohort totaling 2276 patients, including 574 treated with immune checkpoint blockade, we find that mutations in *STK11* and *KEAP1* are associated with poor prognosis across multiple first-line treatment classes.

*Meaning:* Mutations in *STK11* and *KEAP1* are prognostic biomarkers of poor response to both immune checkpoint blockade and chemotherapy.

## Introduction

Advances in personalized medicine have significantly changed clinical decision-making in the treatment of non–small cell lung cancer (NSCLC). Drugs for patients carrying an epidermal growth factor receptor *(EGFR)* or *BRAF* p.V600E mutation, or anaplastic lymphoma kinase *(ALK)* or *ROS1* rearrangements, have significantly improved survival and established the importance of molecularly defined therapies.^1^ However, not all patients with NSCLC have benefited, as a fraction of patients do not carry such actionable mutations. More recently, immunomodulatory cancer drugs such as anti–programmed death-1 (PD-1) have shown significant clinical benefit in NSCLC,^2^ however many patients do not show such benefit, highlighting the need for predictive biomarkers to guide patient stratification strategies.

Measuring tumor programmed death ligand 1 (PD-L1) protein expression using immunohistochemistry assays has been proposed as a rational and biologically sound approach to patient stratification.^3^ Indeed, multiple PD-L1 assays are approved as companion or complementary diagnostics in NSCLC. However, PD-L1 expression alone does not always correlate with response, and additional biomarkers are needed.

Tumor mutational burden (TMB), typically assessed by tallying up all nonsynonymous mutations, has also been explored as an independent predictive biomarker of response to anti–PD-1 treatment.^4^ This measure serves as a proxy for the number of putative neoantigens that could be recognized by immune cells to trigger an immune response.

In addition to markers such as TMB and PD-L1, studies have assessed the role of frequently mutated genes in NSCLC as drivers of primary resistance to immunotherapy. Somatic mutations in serine/threonine kinase 11 (*STK11*)^5-7^ and kelch-like ECH-associated protein 1 (*KEAP1*)^8^ have been proposed as potential modulators of immune response in non-squamous NSCLC. *STK11* has been linked to multiple cellular processes, notably in lipid, glucose, and cholesterol metabolism via activation of 5’ AMP-activated protein kinase (AMPK),^9^ and has been associated with immune escape in a murine model.^10^ *KEAP1* functions as a negative regulator of nuclear factor erythroid 2-related factor 2 (NRF2),^11^ and loss-of-function mutations may contribute to an overactive cytoprotective program. However, genomic data sets from controlled clinical studies are not adequately powered to dissect effects of specific mutations or lack a non–immune checkpoint blockade (ICB) arm to ascertain the predictive nature of those biomarkers. Thus, those studies are challenging to translate into clinical practice to inform treatment options. To assess the predictive or prognostic nature of *STK11* and *KEAP1* mutations in non-squamous NSCLC, we leveraged real-world data from the Flatiron Health Clinico-Genomic Database (CGDB), which includes patients with detailed clinical information and genomic testing by Foundation Medicine.

## Methods

From the CGDB for NSCLC^12^ (April 2019 release; Flatiron Health, New York, NY), we selected patients who had tumor-based genetic testing performed on the FoundationOne CDx or FoundationOne assay (Foundation Medicine Inc, Cambridge, MA). To mitigate the risk of prior treatments affecting results, we focused our analyses on first-line treatment. To ensure that treatment sequencing was correct, we excluded patients with an advanced diagnosis before January 1, 2011, and those who had initiated first-line treatment after 90 days following their advanced diagnosis date. Patients were further selected to have a nonsquamous histology and by their first-line treatment, resulting in 2276 patients across 5 treatment classes (eTable 1, eMethods). We performed time-to-event analysis on real-world, progression-free survival^13^ (rwPFS) and overall survival (OS),^14^ as previously defined.

## Results

Consistent with previous estimates, *STK11* and *KEAP1* mutations were found in 20% (454 of 2276 and 451 of 2276, respectively) of patients and were frequently co-mutated (eFigure 1). Thirty percent (674 of 2276) of patients had tumors which carried either *STK11* and/or *KEAP1* mutations (*STK11*-*KEAP1*) and were enriched for male patients (53.4% vs 42.3%, P < .001, chi-square test), younger age at advanced diagnosis (64.9 vs 66.9 years, P < .001, Student’s t-test), smoking history (96% vs 73.3 %, P < .001, chi-square test), and higher TMB (13.1 vs 7.94 mutations per megabase, P < .001, Student’s t-test) (Table 1). Those results were consistent even when excluding EGFR-mutated patients (eTable 2). KRAS mutations were found in 39% (263 of 674) of *STK11*-*KEAP1* patients (eFigure 1). First-line treatment class was associated with *STK11*-*KEAP1* mutational status, explained by the finding that *EGFR* mutations are mutually exclusive with *STK11*-*KEAP1* and patients carrying EGFR mutations received EGFR tyrosine kinase inhibitors (TKIs) (eFigure 1, Table 1, eTable 3). In a comparison of *STK11*-*KEAP1* vs wild-type patients, excluding EGFR-mutated patients, first-line treatment was not associated with *STK11*-*KEAP1* status (eTable 2).

**Table 1.**
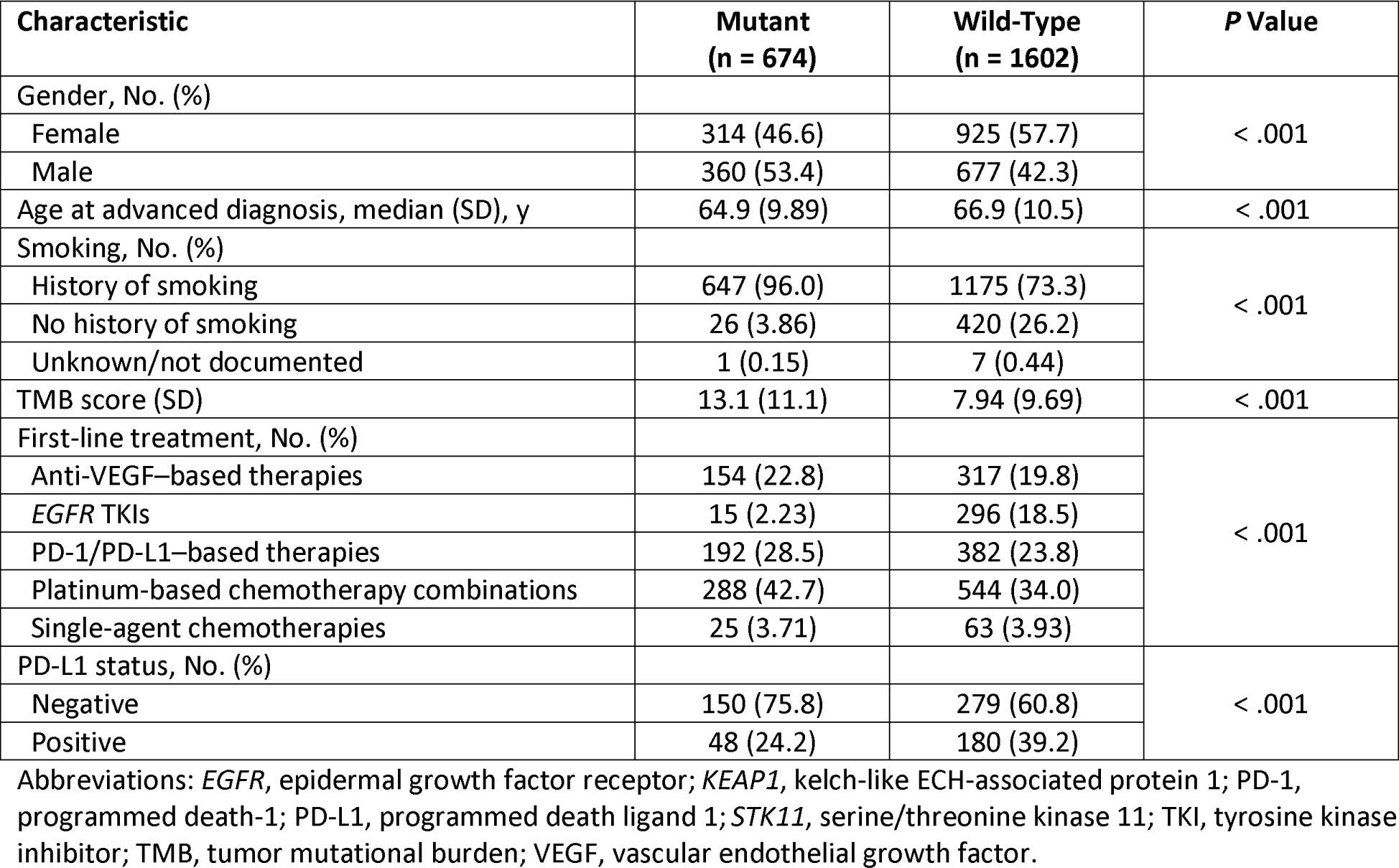
Comparative Table of *STK11*-*KEAP1* Mutated Patients vs Wild-Type Patients.

| Characteristic | Mutant<br>(n = 674) | Wild-Type<br>(n = 1602) | P Value |
| --- | --- | --- | --- |
| Gender, No. (%) |  |  | < .001 |
| Female | 314 (46.6) | 925 (57.7) |  |
| Male | 360 (53.4) | 677 (42.3) |  |
| Age at advanced diagnosis, median (SD), y | 64.9 (9.89) | 66.9 (10.5) | < .001 |
| Smoking, No. (%) |  |  | < .001 |
| History of smoking | 647 (96.0) | 1175 (73.3) |  |
| No history of smoking | 26 (3.86) | 420 (26.2) |  |
| Unknown/not documented | 1 (0.15) | 7 (0.44) |  |
| TMB score (SD) | 13.1 (11.1) | 7.94 (9.69) | < .001 |
| First-line treatment, No. (%) |  |  | < .001 |
| Anti-VEGF–based therapies | 154 (22.8) | 317 (19.8) |  |
| *EGFR* TKIs | 15 (2.23) | 296 (18.5) |  |
| PD-1/PD-L1–based therapies | 192 (28.5) | 382 (23.8) |  |
| Platinum-based chemotherapy combinations | 288 (42.7) | 544 (34.0) |  |
| Single-agent chemotherapies | 25 (3.71) | 63 (3.93) |  |
| PD-L1 status, No. (%) |  |  | < .001 |
| Negative | 150 (75.8) | 279 (60.8) |  |
| Positive | 48 (24.2) | 180 (39.2) |  |

Abbreviations: *EGFR*, epidermal growth factor receptor; *KEAP1*, kelch-like ECH-associated protein 1; PD-1, programmed death-1; PD-L1, programmed death ligand 1; *STK11*, serine/threonine kinase 11; TKI, tyrosine kinase inhibitor; TMB, tumor mutational burden; VEGF, vascular endothelial growth factor.

*STK11* mutations were previously reported to be associated with low levels of T-cell inflammation and tumor PD-L1 expression.^6^ Consistent with previous reports, patients with *STK11*-*KEAP1* mutations were enriched for negative PD-L1 staining (75.8% vs 60.8%, P < .001, chi-square test; Table 1), as were patients with EGFR mutations (eTable 3).

We performed multivariate Cox proportional hazards modeling, including age at advanced diagnosis, gender, TMB (continuous variable), and *STK11*-*KEAP1* mutational status for rwPFS for each treatment class independently. *STK11* and *KEAP1* mutations were both associated with poor prognosis for rwPFS across treatment classes (Figure 1A). We then focused on anti–PD-1/PD-L1 and chemotherapy treatment classes and tested whether *STK11*-*KEAP1* mutations showed a treatment-specific effect by including an interaction term (*STK11*-*KEAP1* * treatment) in our previous Cox model. Consistent with the previous model, *STK11* and *KEAP1* mutations were prognostic and did not show different treatment-specific effects (Figure 1B).

**Figure 1.**
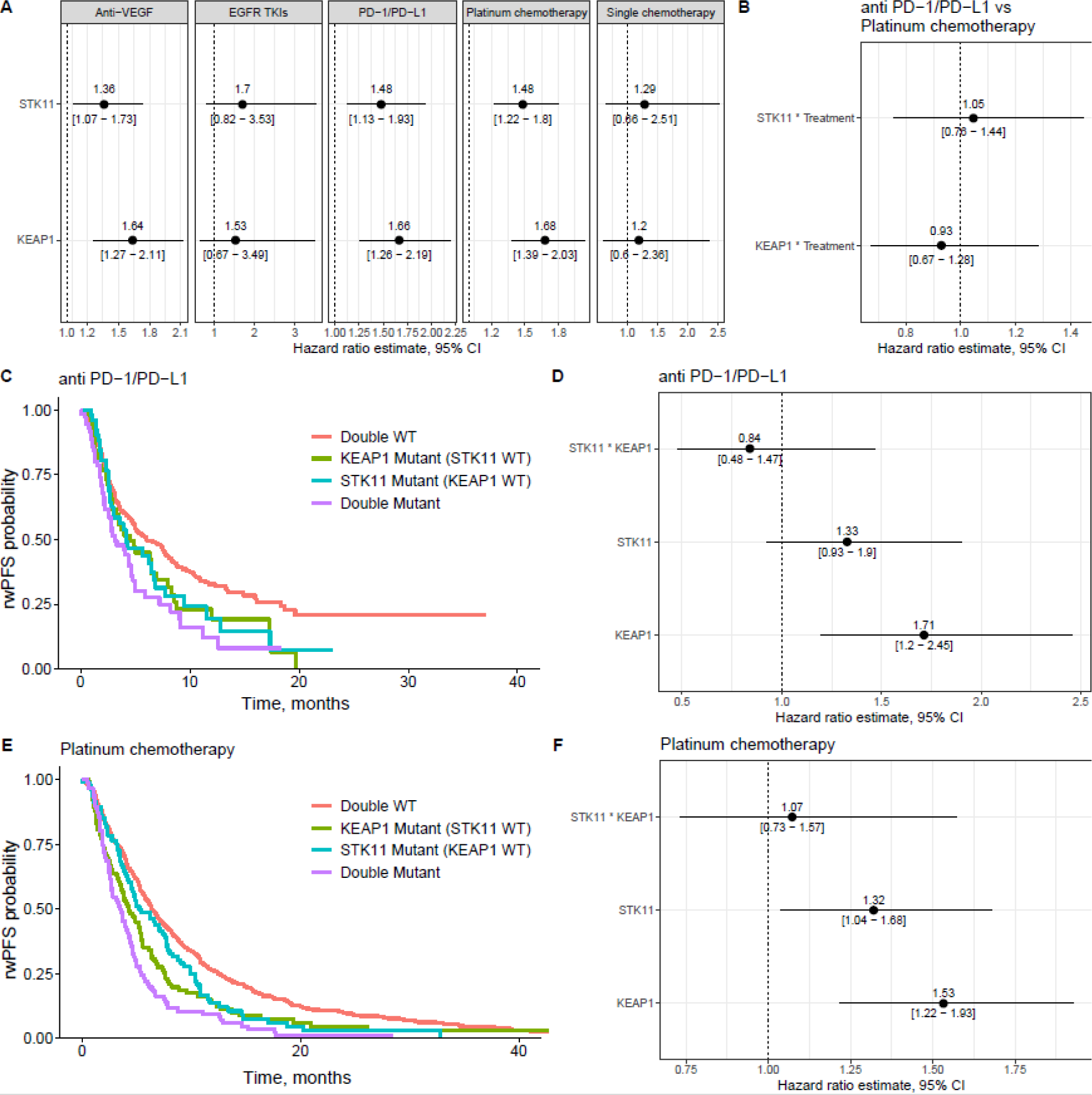
Effect of *STK11* and *KEAP1* Somatic Mutations on rwPFS in a First-Line Setting. (A) Forest plot of the hazard ratios of mutations in *STK11* or *KEAP1* across different treatment classes. (B) Forest plot of the hazard ratios of the interaction terms of *STK11* or *KEAP1* mutations and treatment (platinum chemotherapy vs PD-1/PD-L1). (C) Kaplan-Meier curves of PD-1/PD-L1–treated patients according to *STK11*-*KEAP1* status. (D) Forest plot of the hazard ratios of *STK11* and *KEAP1* in PD-1/PD-L1–treated patients. (E) Kaplan-Meier curves of platinum chemotherapy–treated patients according to *STK11*-*KEAP1* status. (F) Forest plot of the hazard ratios of *STK11* and *KEAP1* in platinum chemotherapy–treated patients. *EGFR*, epidermal growth factor receptor; *KEAP1*, kelch-like ECH-associated protein 1; PD-1, programmed death-1; PD-L1, programmed death ligand 1; rwPFS, real-world progression-free survival; *STK11*, serine/threonine kinase 11; TKI, tyrosine kinase inhibitor; VEGF, vascular endothelial growth factor; WT, wild-type.

We tested the independent contributions of *STK11* and *KEAP1* mutations to poor prognosis by testing them in a multivariate model including the interaction between mutations in the two genes. Both genes were associated with lower rwPFS, and *KEAP1*-only patients fared worse than patients with *STK11*-only mutations, while patients with double-mutational status had the worst outcomes (Figure 1C-F). The interaction term was not associated with rwPFS, suggesting that *STK11* and *KEAP1* mutations have an additive effect. Those results were consistent across anti–PD-1/PD-L1 and chemotherapy treatment classes (Figure 1C-F).

We then performed an analogous analysis using OS as the endpoint and observed the prognostic nature of *STK11*-*KEAP1* mutations to be highly consistent with observations for rwPFS (Figure 2A-F).

**Figure 2.**
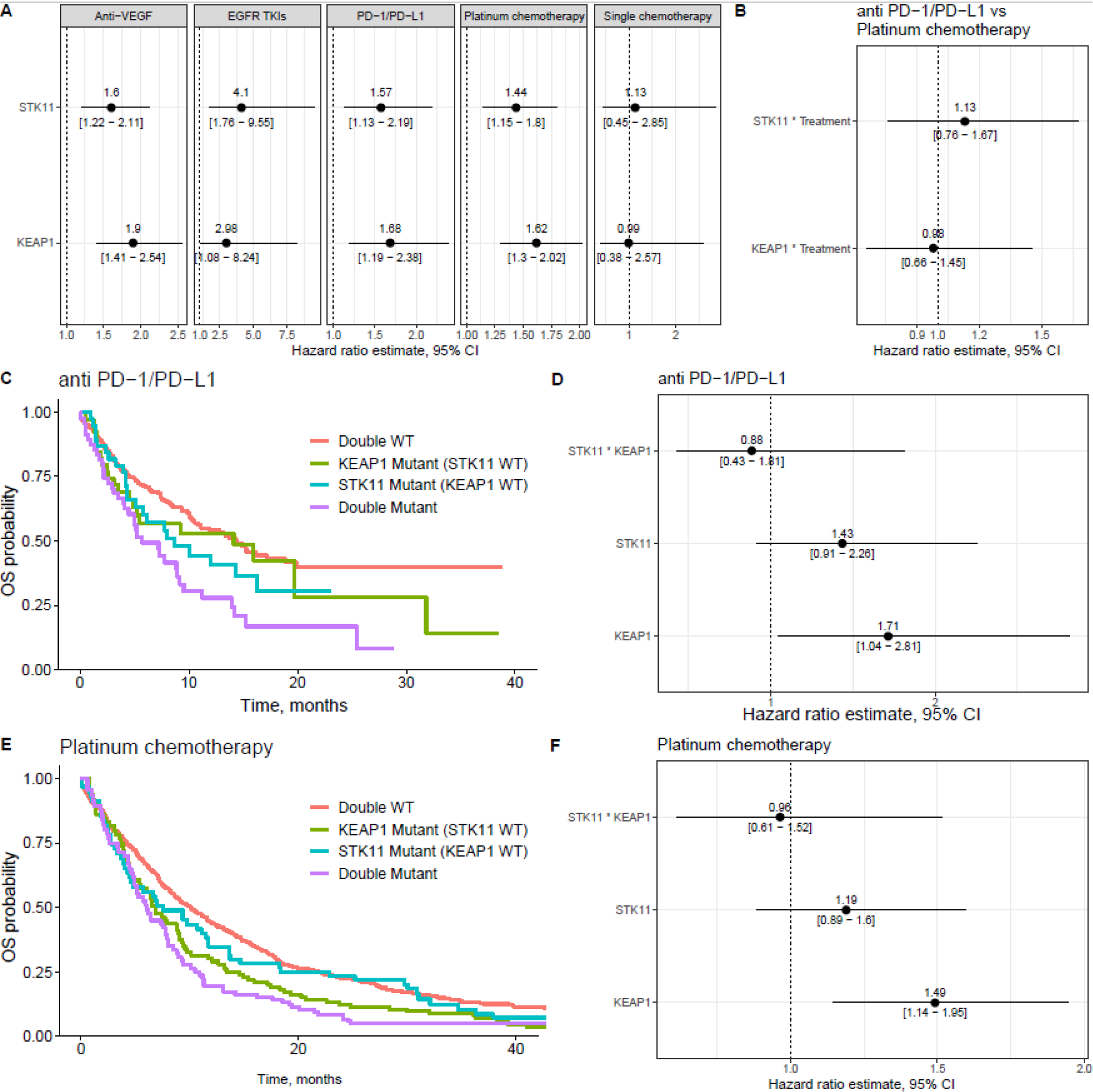
Effect of *STK11* and *KEAP1* Somatic Mutations on OS in a First-Line Setting. (A) Forest plot of the hazard ratios of mutations in *STK11* or *KEAP1* across different treatment classes. (B) Forest plot of the hazard ratios of the interaction terms of *STK11* or *KEAP1* mutations and treatment (platinum chemotherapy vs PD-1/PD-L1). (C) Kaplan-Meier curves of PD-1/PD-L1–treated patients according to *STK11*-*KEAP1* status. (D) Forest plot of the hazard ratios of *STK11* and *KEAP1* in PD-1/PD-L1–treated patients. (E) Kaplan-Meier curves of platinum chemotherapy–treated patients according to *STK11*-*KEAP1* status. (F) Forest plot of the hazard ratios of *STK11* and *KEAP1* in platinum chemotherapy–treated patients. *EGFR*, epidermal growth factor receptor; *KEAP1*, kelch-like ECH-associated protein 1; OS, overall survival; PD-1, programmed death-1; PD-L1, programmed death ligand 1; *STK11*, serine/threonine kinase 11; TKI, tyrosine kinase inhibitor; VEGF, vascular endothelial growth factor; WT, wild-type.

We performed the analyses described above in *KRAS*-mutated patients to test the utility of *STK11*-*KEAP1* mutations as a predictive biomarker for this patient population (eFigure 2). *STK11*-*KEAP1* mutations were associated with poor prognosis in this patient subset in both anti–PD-1/PD-L1- and chemotherapy-treated populations, consistent with our overall findings.

## Discussion

Our comprehensive profiling of *STK11* and *KEAP1* mutations in nonsquamous NSCLC demonstrated that these mutations confer a poor prognosis, regardless of treatment class. Our results complement previous reports suggesting that *STK11*-*KEAP1* patients respond poorly to ICB by highlighting that this effect is observed across all treatment classes and is not specific to ICB cohorts.^5,6,8^

## Limitations

Real-world data are retrospective and observational and thus may not offer the same robustness as prospective randomized clinical trials. Factors that influence clinical decision-making but are not explicitly captured by real-world data sets may exist and thus confound analyses. Other factors such as tumor evolutionary dynamics between specimen collection, diagnosis, and treatment start or during treatment may influence the associations. Moreover, although the cohort was large, it might not be sufficiently powered to capture a low-effect-size interaction between *STK11* and *KEAP1* mutations.

## Conclusion

Our results provide evidence against previous reports suggesting that *STK11*-*KEAP1* mutations are predictive biomarkers for anti–PD-1/PD-L1 therapy.^5,6,8^ Using a cohort of 2276 patients with NSCLC, we show that *STK11* and *KEAP1* mutations are associated with poor prognosis across all therapy classes and should not be used as a patient selection marker for ICB.

## Data Availability

Data was licensed from Flatiron Health.

## Acknowledgements

Contribution of authors: SP-C and AMW performed analyses. All authors participated in writing the article. Editorial assistance was provided by Michelle Utton-Mishra, PhD, and Jay Rathi, MA, of Spark Medica Inc, according to Good Publication Practice guidelines, funded by Bristol-Myers Squibb. All authors are BMS employees and shareholders. PD is a J&J shareholder, RD is a Merck and J&J shareholder.

## eMethods

### Treatment Consolidation

First-line treatment data were aggregated in 5 broad treatment classes according to Flatiron Health rules. In summary, regimens that contained anti–PD-1 or anti–PD-L1 were considered “PD-1/PD-L1–based therapies”, those that contained *EGFR* TKIs as “*EGFR* TKIs”, and those that contained anti-VEGF as “anti-VEGF–based therapies”. Regimens including a platinum-based and any other chemotherapeutic agent, but not drugs from the above-mentioned class, were classified as “platinum-based chemotherapy combinations”. Regimens with a single chemotherapeutic agent were considered “single-agent chemotherapies”.

### Genomic Assay

We selected patient specimens profiled on bait sets DX1, T4b, T5a, or T7 of the Foundation Medicine FoundationOne CDx or FoundationOne assay, as they are performed on tumor material (as opposed to blood) and they contain tumor protein p53 (*TP53), STK11, KEAP1*, and *KRAS* in their gene panel.

### *STK11, KEAP1*, and *TP53* Mutations

We aggregated gene-specific alterations, filtering for missense mutations, truncations, and deletions. For OncoPrint visual display, all mutations were considered. For Cox proportional hazards and Kaplan-Meier modeling, patients were labeled as mutant for a gene if they had at least 1 qualifying (missense, truncation, and deletion) mutation in that gene. OncoPrint plots were created using ComplexHeatmap R package.^15^

### *KRAS* Mutations

For simplicity, we labeled only patients who carried a missense mutation in the hotspot locus G12-13 as *KRAS*-mutated, whereas patients carrying other alterations were considered as *KRAS*–wild type.

### Time-to-Event Analysis

We performed rwPFS and OS analysis using Cox proportional-hazards modeling in R. Kaplan-Meier curves were displayed using the R package survminer.

### PD-L1 Harmonization

To obtain PD-L1 immunohistochemistry data for as many patients as possible, we harmonized the PD-L1 immunohistochemistry data from different tests by considering numerical values >50% tumor cell scoring as positive.

## Supplementary Tables and Figures

**eTable 1.** Cohort Description.

| Characteristic | All Patients<br>(N = 2276) | No. With Data |
| --- | --- | --- |
| Gender, No. (%) |  | 2276 |
| Female | 1239 (54.4) |  |
| Male | 1037 (45.6) |  |
| Age at advanced diagnosis, median (SD), y | 66.3 (10.4) | 2276 |
| Smoking, No. (%) |  | 2276 |
| History of smoking | 1822 (80.1) |  |
| No history of smoking | 446 (19.6) |  |
| Unknown/not documented | 8 (0.35) |  |
| TMB score (SD) | 9.47 (10.4) | 2261 |
| First-line treatment, No. (%) |  | 2276 |
| Anti-VEGF-based therapies | 471 (20.7) |  |
| *EGFR* TKIs | 311 (13.7) |  |
| PD-1/PD-L1-based therapies | 574 (25.2) |  |
| Platinum-based chemotherapy combinations | 832 (36.6) |  |
| Single-agent chemotherapies | 88 (3.87) |  |
| PD-L1 status, No. (%) |  | 657 |
| Negative | 429 (65.3) |  |
| Positive | 228 (34.7) |  |

Abbreviations: *EGFR*, epidermal growth factor receptor; PD-1, programmed death-1; PD-L1, programmed death ligand 1; TKI, tyrosine kinase inhibitor; TMB, tumor mutational burden; VEGF, vascular endothelial growth factor.

**eTable 2.**
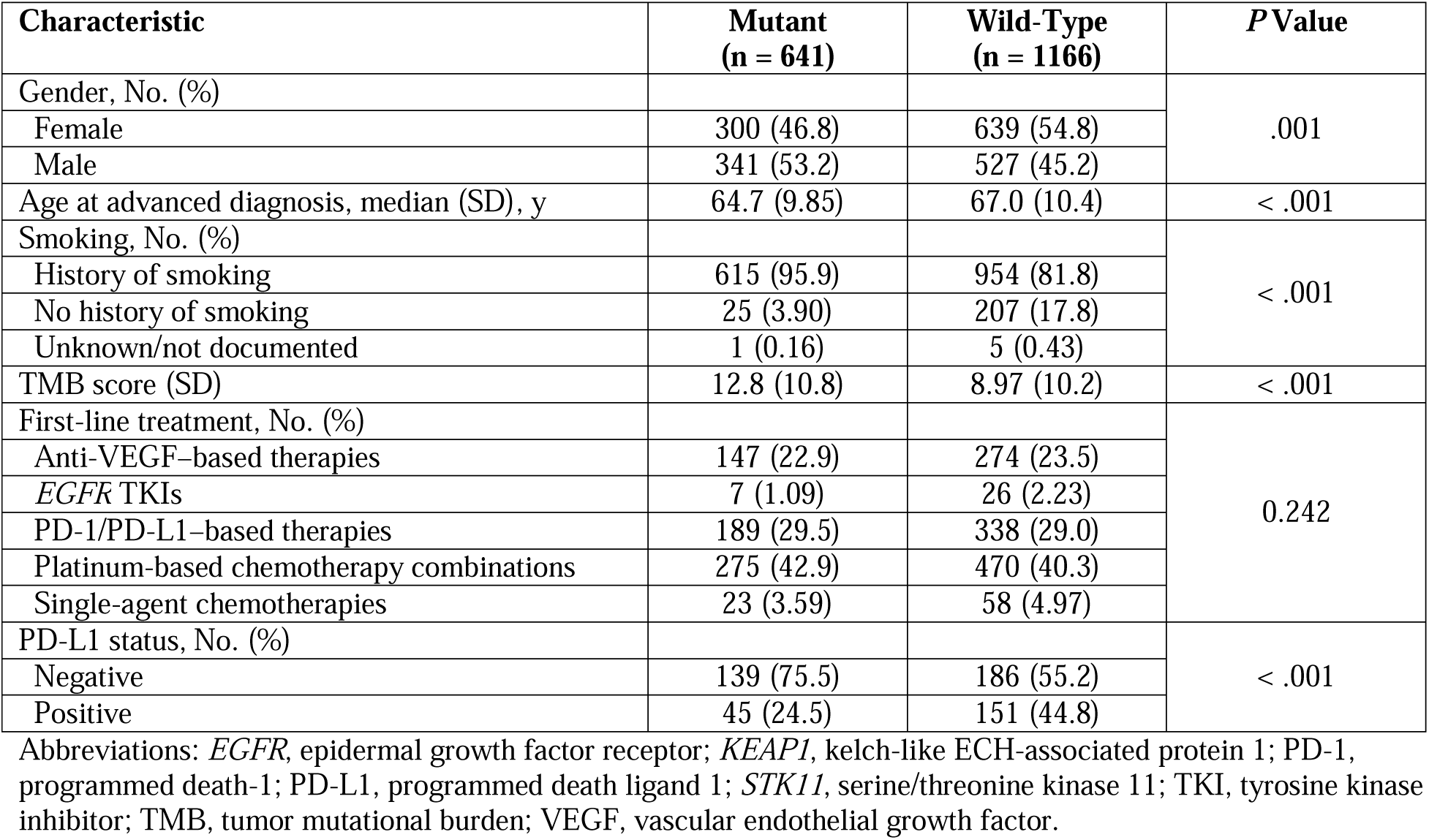
Comparative Table of STK11-KEAP1 Mutated Patients vs Wild-Type Patients, Excluding EGFR-Mutated Patients.

| Characteristic | Mutant<br>(n = 641) | Wild-Type<br>(n = 1166) | P Value |
| --- | --- | --- | --- |
| Gender, No. (%) |  |  | .001 |
| Female | 300 (46.8) | 639 (54.8) |  |
| Male | 341 (53.2) | 527 (45.2) |  |
| Age at advanced diagnosis, median (SD), y | 64.7 (9.85) | 67.0 (10.4) | < .001 |
| Smoking, No. (%) |  |  | < .001 |
| History of smoking | 615 (95.9) | 954 (81.8) |  |
| No history of smoking | 25 (3.90) | 207 (17.8) |  |
| Unknown/not documented | 1 (0.16) | 5 (0.43) |  |
| TMB score (SD) | 12.8 (10.8) | 8.97 (10.2) | < .001 |
| First-line treatment, No. (%) |  |  | 0.242 |
| Anti-VEGF-based therapies | 147 (22.9) | 274 (23.5) |  |
| EGFR TKIs | 7 (1.09) | 26 (2.23) |  |
| PD-1/PD-L1-based therapies | 189 (29.5) | 338 (29.0) |  |
| Platinum-based chemotherapy combinations | 275 (42.9) | 470 (40.3) |  |
| Single-agent chemotherapies | 23 (3.59) | 58 (4.97) |  |
| PD-L1 status, No. (%) |  |  | < .001 |
| Negative | 139 (75.5) | 186 (55.2) |  |
| Positive | 45 (24.5) | 151 (44.8) |  |

Abbreviations: *EGFR*, epidermal growth factor receptor; *KEAP1*, kelch-like ECH-associated protein 1; PD-1, programmed death-1; PD-L1, programmed death ligand 1; *STK11*, serine/threonine kinase 11; TKI, tyrosine kinase inhibitor; TMB, tumor mutational burden; VEGF, vascular endothelial growth factor.

**eTable 3.**
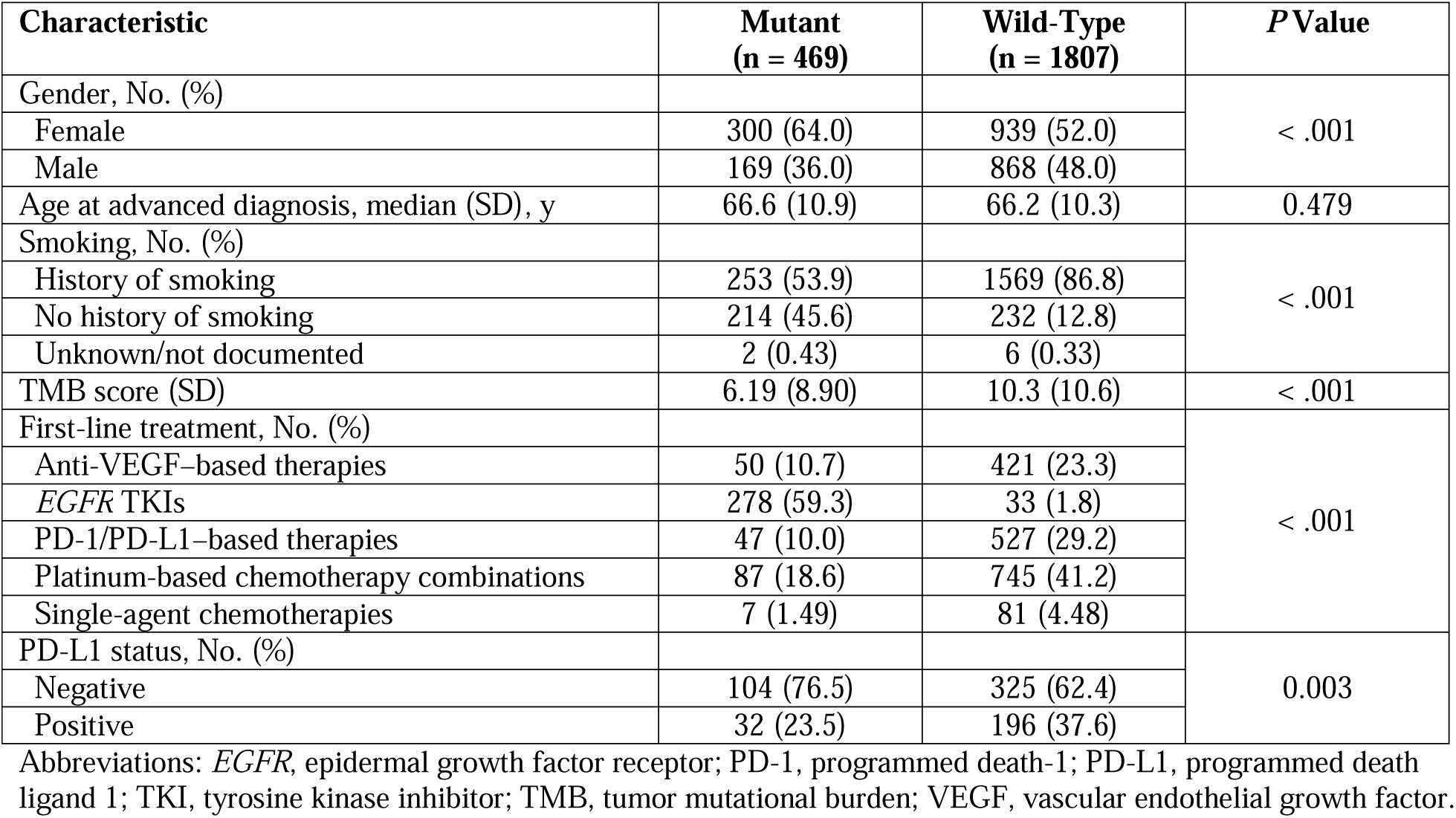
Comparison of EGFR Mutant vs EGFR Wild-Type Patients.

| Characteristic | Mutant<br>(n = 469) | Wild-Type<br>(n = 1807) | P Value |
| --- | --- | --- | --- |
| Gender, No. (%) |  |  | < .001 |
| Female | 300 (64.0) | 939 (52.0) |  |
| Male | 169 (36.0) | 868 (48.0) |  |
| Age at advanced diagnosis, median (SD), y | 66.6 (10.9) | 66.2 (10.3) | 0.479 |
| Smoking, No. (%) |  |  | < .001 |
| History of smoking | 253 (53.9) | 1569 (86.8) |  |
| No history of smoking | 214 (45.6) | 232 (12.8) |  |
| Unknown/not documented | 2 (0.43) | 6 (0.33) |  |
| TMB score (SD) | 6.19 (8.90) | 10.3 (10.6) | < .001 |
| First-line treatment, No. (%) |  |  | < .001 |
| Anti-VEGF-based therapies | 50 (10.7) | 421 (23.3) |  |
| EGFR TKIs | 278 (59.3) | 33 (1.8) |  |
| PD-1/PD-L1-based therapies | 47 (10.0) | 527 (29.2) |  |
| Platinum-based chemotherapy combinations | 87 (18.6) | 745 (41.2) |  |
| Single-agent chemotherapies | 7 (1.49) | 81 (4.48) |  |
| PD-L1 status, No. (%) |  |  | 0.003 |
| Negative | 104 (76.5) | 325 (62.4) |  |
| Positive | 32 (23.5) | 196 (37.6) |  |

Abbreviations: *EGFR*, epidermal growth factor receptor; PD-1, programmed death-1; PD-L1, programmed death ligand 1; TKI, tyrosine kinase inhibitor; TMB, tumor mutational burden; VEGF, vascular endothelial growth factor.

**eFigure 1.**
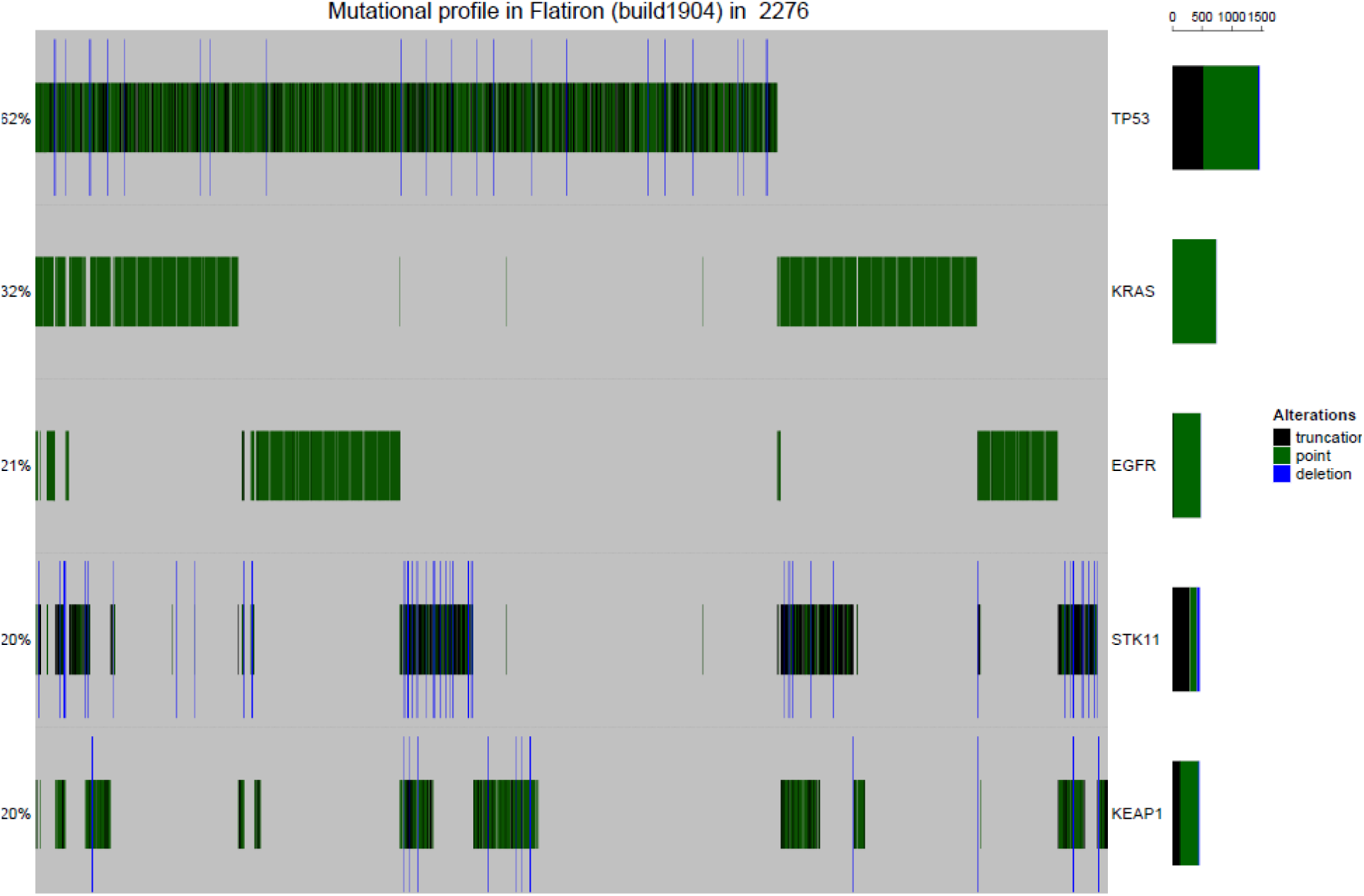
OncoPrint of Prevalent Mutations in 2276 Patients With NSCLC Used for This Analysis. Abbreviations: *EGFR*, epidermal growth factor receptor; *KEAP1*, kelch-like ECH-associated protein 1; NSCLC, non–small cell lung cancer; *STK11*, serine/threonine kinase 11; *TP53*, tumor protein p53.

**eFigure 2.**
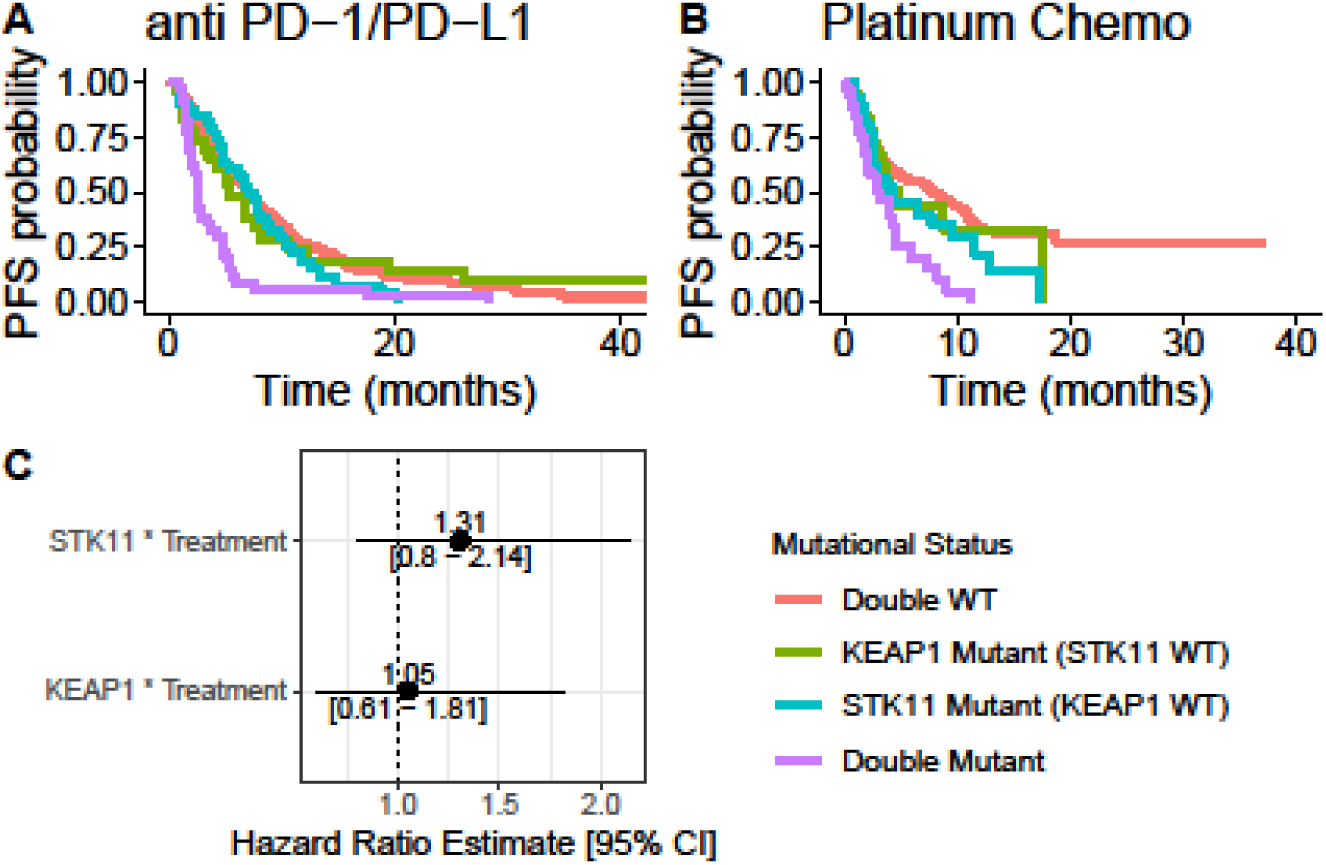
Effect of STK11 and KEAP1 Somatic Mutations on rwPFS in a First-Line Setting in KRAS-Mutated Patients. (A) Kaplan-Meier curves of PD-1/PD-L1–treated patients according to *STK11-KEAP1* status. (B) Kaplan-Meier curves of platinum chemotherapy–treated patients according to *STK11-KEAP1* status. (C) Forest plot of the hazard ratios of the interaction terms of *STK11* or *KEAP1* mutations and treatment (platinum chemotherapy vs PD-1/PD-L1). Abbreviations: *KEAP1*, kelch-like ECH-associated protein 1; PD-1, programmed death-1; PD-L1, programmed death ligand 1; rwPFS, real-world progression-free survival; *STK11*, serine/threonine kinase 11; WT, wild-type.

